# Determinants of severe adverse outcomes among low-birth-weight neonates admitted to a county referral hospital in Kenya: a mixed-methods cross-sectional study

**DOI:** 10.64898/2026.07.20.26358516

**Authors:** Judy Cheptoo, Morris Senghor Shisanya, Mukhtar Kiprono, Everlyne Nyanchera

**Author notes:** Corresponding author. (MSS);.

## Abstract

**Background:** Low-birth-weight (LBW) neonates carry a disproportionate burden of morbidity and mortality in resource-limited newborn units, yet facility-level evidence on the determinants of severe in-hospital outcomes at county referral level in Kenya is limited. We determined the maternal, neonatal and care-related determinants of severe adverse outcomes among LBW neonates admitted to Kericho County Referral Hospital (KCRH) and used healthcare-provider perspectives to explain the quantitative findings.

**Methods:** We conducted a facility-based, convergent mixed-methods cross-sectional study. Quantitative data were obtained from 169 LBW neonate–mother pairs through structured maternal interviews and clinical-record abstraction; qualitative data came from nine key informant interviews with newborn-unit healthcare workers. A severe adverse outcome was defined as the occurrence of at least one of respiratory distress, sepsis, hypothermia, hypoglycaemia, prolonged neonatal unit stay (≥7 days) or neonatal death. Associations were examined using bivariate tests and a multivariable binary logistic regression model entering 13 candidate predictors simultaneously, with multicollinearity, calibration and discrimination diagnostics. Interviews were analysed thematically and integrated with the quantitative results in a joint display.

**Results:** A severe adverse outcome occurred in 136 of 169 neonates (80.5%); respiratory distress was the most common single complication (69.8%), and neonates experienced a mean of 2.46 (SD 1.31) adverse outcomes. In the adjusted model (omnibus χ²(13) = 57.70, p < 0.001; Nagelkerke R² = 0.46; area under the receiver-operating-characteristic curve = 0.879), three factors independently predicted severe adverse outcome: lower birth weight (adjusted odds ratio [AOR] 0.997 per gram, 95% CI 0.995–0.999, p = 0.012), maternal pregnancy-induced hypertension (AOR 18.49, 95% CI 1.87–182.47, p = 0.013) and warm-chain care (AOR 10.94, 95% CI 1.40–85.41, p = 0.023; a direction consistent with confounding by indication). Providers emphasised the fragile first hour of stabilisation, staffing and workload, warm-chain maintenance, infection prevention, commodity availability and referral coordination.

**Conclusions:** Severe adverse outcomes were near-universal among LBW neonates at KCRH and were driven by neonatal biological vulnerability, maternal hypertensive disease and the readiness of newborn-unit care. Improving outcomes requires early, birth-weight-based risk stratification of the smallest neonates alongside strengthening of antenatal detection of hypertension and reliable, timely newborn-unit care processes.

## Introduction

Low birth weight (LBW), defined as a birth weight below 2,500 g, remains a major global public health concern because of its strong association with neonatal morbidity, mortality and long-term developmental impairment [1–3]. An estimated 20 million LBW infants are born each year, and both the prevalence and the associated risk of complications are concentrated in low- and middle-income countries, particularly in sub-Saharan Africa and South Asia [4–7].

LBW neonates are especially susceptible to respiratory distress, neonatal sepsis, hypothermia, hypoglycaemia and feeding difficulties, each of which increases the risk of prolonged hospitalisation and early death [8–11]. These complications arise from physiological immaturity—limited thermoregulation, reduced glycogen and fat stores, immature lungs and immune function—and often occur together, so that a single neonate may experience several overlapping morbidities [6, 12].

In Kenya, the pattern mirrors global trends. The 2022 Kenya Demographic and Health Survey reported a neonatal mortality rate of 22 deaths per 1,000 live births, with preterm birth and LBW among the leading contributors [13, 14]. However, most research in the region has focused on the antenatal and social causes of LBW rather than on what determines outcomes after LBW neonates are admitted to hospital newborn units [15–17]. Consequently, facility-level evidence on the maternal, neonatal and care-related factors that shape in-hospital morbidity and mortality among this vulnerable group remains scarce, particularly in county referral settings.

Kericho County Referral Hospital (KCRH) is the principal referral facility in Kericho County and admits a substantial number of LBW neonates, many of whom present with serious complications while the unit contends with constraints in staffing, equipment and commodities. At this level of care, neonatal biological vulnerability and health-system readiness intersect. Two complementary frameworks help to structure this intersection: the Social Ecological Model, which locates health outcomes within interacting individual, interpersonal, community, institutional and policy levels [18, 19]; and the Three Delays Model, which identifies delays in deciding to seek care, reaching care and receiving adequate care as critical, modifiable points along the pathway to survival [20–22].

Against this background, this study aimed to determine the maternal, neonatal and care-related determinants of severe adverse outcomes among LBW neonates admitted to the newborn unit at KCRH, and to use healthcare-provider perspectives to explain the quantitative findings. We addressed the question: which maternal, neonatal and care-related factors are associated with severe adverse outcomes among LBW neonates, and how do providers explain these outcomes within the newborn-unit care context?

## Materials and methods

### Study design and reporting

We conducted a facility-based, convergent mixed-methods cross-sectional study integrating a quantitative strand (clinical and maternal data) with a qualitative strand (provider interviews). The quantitative component measured the burden of adverse outcomes and their maternal, neonatal and care-related determinants, while the qualitative component explained the care processes and system conditions influencing those outcomes. Reporting follows the Strengthening the Reporting of Observational Studies in Epidemiology (STROBE) guideline for the quantitative strand [23] and the Consolidated Criteria for Reporting Qualitative Research (COREQ) for the qualitative strand [24].

### Study setting

The study was conducted in the newborn unit of KCRH, a public county referral hospital in Kericho County, Kenya. The unit serves as a regional referral centre for high-risk neonates and manages a high volume of LBW admissions, receiving both inborn neonates and neonates referred from lower-level facilities across the county.

### Study population and eligibility

The quantitative population comprised LBW neonates (birth weight < 2,500 g) admitted to the newborn unit during the study period, together with their biological mothers. Neonates were eligible if they had a documented birth weight below 2,500 g and complete clinical records including birth weight, gestational age, Apgar score and final admission outcome. Neonates were excluded if essential documentation (gestational age or final admission outcome) was missing, or if they were transferred to another facility before outcome determination. Mothers were eligible if they were aged 18 years or older (or emancipated minors), reachable during admission and able to give written informed consent; mothers who were critically ill and unable to consent were excluded. For the qualitative strand, healthcare providers (pediatricians, nurses, clinical officers and the unit manager) with at least six months of newborn-unit experience and direct involvement in LBW care were eligible.

### Sample size and sampling

The quantitative sample size was calculated using Cochran’s formula for proportions with a finite population correction, assuming a 95% confidence level, a 5% margin of error, a maximum-variability outcome proportion of 0.5 and an average 12-week admission volume of approximately 173 LBW neonates. This yielded 120 pairs, increased to 134 after a 10% non-response adjustment. Neonate–mother pairs were enrolled consecutively as they were admitted; the final cleaned analytic dataset comprised 169 complete pairs. Healthcare providers were purposively sampled until thematic saturation, resulting in nine key informant interviews.

### Data collection

Data collection commenced on 3^rd^ April 2026 and ended on 7^th^ June 2026. Quantitative data were collected by trained research assistants using a structured maternal questionnaire and a clinical-record abstraction form. The questionnaire captured sociodemographic and contextual characteristics, obstetric and antenatal history, care-seeking and referral information, and maternal clinical conditions; the abstraction form captured neonatal characteristics (birth weight, gestational age, Apgar score, sex, mode of delivery, congenital anomalies), care-related variables (time to initiation of care, admission status, skilled-personnel availability, warm-chain maintenance, essential drug or feed availability, referral handling) and documented in-hospital outcomes. Instruments were developed from the study’s conceptual framework and reviewed literature, pretested at a comparable facility, and refined for clarity. Qualitative data were collected by the principal investigator through in-depth key informant interviews using a semi-structured guide that explored provider perspectives on determinants of adverse outcomes, care-related practices, facility-level challenges, resource availability and referral processes. Interviews lasted approximately 30–45 minutes, were audio-recorded with consent, supplemented with field notes and transcribed verbatim.

### Outcome and predictor variables

The primary outcome was a binary severe adverse neonatal outcome, defined as the occurrence during admission of at least one of respiratory distress, neonatal sepsis, hypothermia, hypoglycaemia, prolonged neonatal-unit stay of seven days or more, or neonatal death. A broader “any adverse outcome” indicator was examined descriptively but not used for regression, because only nine neonates recorded no adverse outcome, producing an extremely imbalanced variable unsuitable for multivariable modelling. Candidate predictors, guided by the conceptual framework, spanned neonatal factors (birth weight, gestational age, preterm birth, Apgar score, congenital anomalies and a neonatal risk-factor count), maternal factors (pregnancy-induced hypertension [PIH], maternal age, parity and antenatal-care attendance) and care-related factors (skilled-personnel availability, warm-chain maintenance and referral status).

### Quantitative analysis

Data were analysed in IBM SPSS Statistics version 26. Categorical variables were summarised as frequencies and percentages and continuous variables as means and standard deviations. Bivariate associations with severe adverse outcome were tested using the Pearson chi-square test (or Fisher’s exact test where expected cell counts were small) for categorical variables and independent-samples t tests for continuous variables, with Mann–Whitney U tests used to confirm concordance; crude odds ratios with 95% confidence intervals were obtained from univariate logistic regression. Before multivariable modelling, multicollinearity among candidate predictors was assessed using tolerance and the variance inflation factor (VIF), with VIF < 5 considered acceptable. A multivariable binary logistic regression model was then fitted, entering all 13 candidate predictors simultaneously (Enter method). Model performance was assessed using the omnibus test of model coefficients, the Hosmer–Lemeshow goodness-of-fit test, Nagelkerke R², the overall classification accuracy and the area under the receiver-operating-characteristic (ROC) curve. Statistical significance was set at p < 0.05.

### Qualitative analysis and integration

Interview transcripts were analysed thematically following an established six-phase approach—familiarisation, coding, theme development, review, definition and reporting—supported by NVivo software [25]. Coding was initially guided by the interview domains and the study’s care-related and delay constructs, while remaining open to emergent subthemes. The quantitative and qualitative findings were then integrated in a joint display that aligned the key quantitative result for each objective with the supporting qualitative theme and a combined interpretation.

### Ethical considerations

Ethical approval was obtained from the Kabarak University Institutional Scientific and Ethics Review Committee and the National Commission for Science, Technology and Innovation (NACOSTI), with administrative authorisation from the Kericho County health management and KCRH. Written informed consent was obtained from all mothers and healthcare providers before data collection; for mothers unable to read or write, the information was read aloud and consent indicated by thumbprint in the presence of an independent literate witness. Participation was voluntary and could be withdrawn at any time without affecting care. Each participant was assigned a unique identifier, no personal identifiers were recorded on data-collection tools, hard-copy records were stored under lock and electronic files were password-protected, with audio recordings encrypted and accessible only to the research team.

## Results

### Participant characteristics

The analysis included 169 LBW neonate–mother pairs. The cohort consisted entirely of low-birth-weight neonates, of whom 18.3% were very low birth weight (< 1,500 g) and most (85.8%) were preterm; the mean birth weight was 1,784.5 g (SD 389.7) and the mean gestational age 32.4 weeks (SD 3.9). Antenatal-care attendance was high (89.3%), and maternal pregnancy-induced hypertension was recorded in 25.4% of mothers. The sex distribution was near-equal (49.7% male), and congenital anomalies were documented in 16.6% of neonates. Sample characteristics are summarised in Table 1.

**Table 1.**
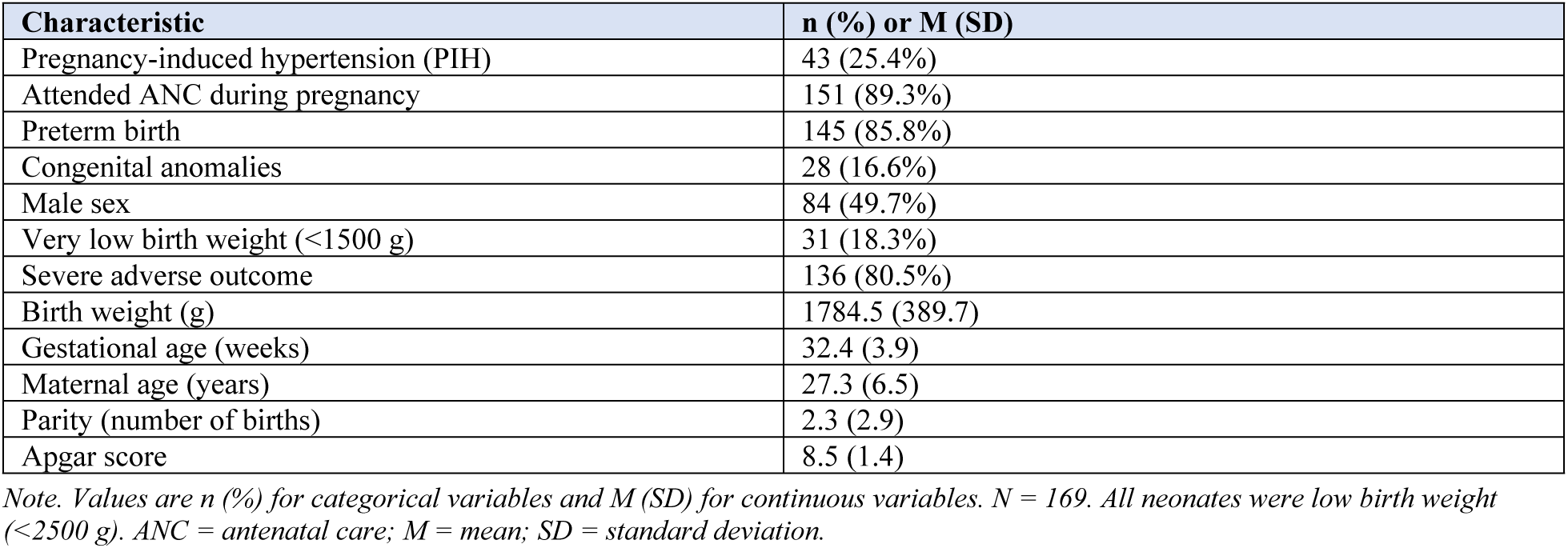
Sample characteristics of neonates and mothers (N = 169).

| Characteristic | n (%) or M (SD) |
| --- | --- |
| Pregnancy-induced hypertension (PIH) | 43 (25.4%) |
| Attended ANC during pregnancy | 151 (89.3%) |
| Preterm birth | 145 (85.8%) |
| Congenital anomalies | 28 (16.6%) |
| Male sex | 84 (49.7%) |
| Very low birth weight (<1500 g) | 31 (18.3%) |
| Severe adverse outcome | 136 (80.5%) |
| Birth weight (g) | 1784.5 (389.7) |
| Gestational age (weeks) | 32.4 (3.9) |
| Maternal age (years) | 27.3 (6.5) |
| Parity (number of births) | 2.3 (2.9) |
| Apgar score | 8.5 (1.4) |

*Note. Values are n (%) for categorical variables and M (SD) for continuous variables. N = 169. All neonates were low birth weight (<2500 g). ANC = antenatal care; M = mean; SD = standard deviation.*

### Types and burden of adverse outcomes

Adverse outcomes were near-universal. Overall, 160 neonates (94.7%) experienced at least one adverse outcome, and 136 (80.5%) experienced a severe adverse outcome, with a mean of 2.46 (SD 1.31) outcomes per neonate. The most common single complication was respiratory distress (118 neonates; 69.8%), followed by prolonged neonatal-unit stay of seven days or more (81; 47.9%), hypoglycaemia (75; 44.4%), sepsis (65; 38.5%) and hypothermia (60; 35.5%); low Apgar score was recorded in 15 neonates (8.9%), neonatal death in 12 (7.1%) and developmental delay in 4 (2.4%). The distribution of specific adverse outcomes is shown in Fig 1.

**Fig 1.**
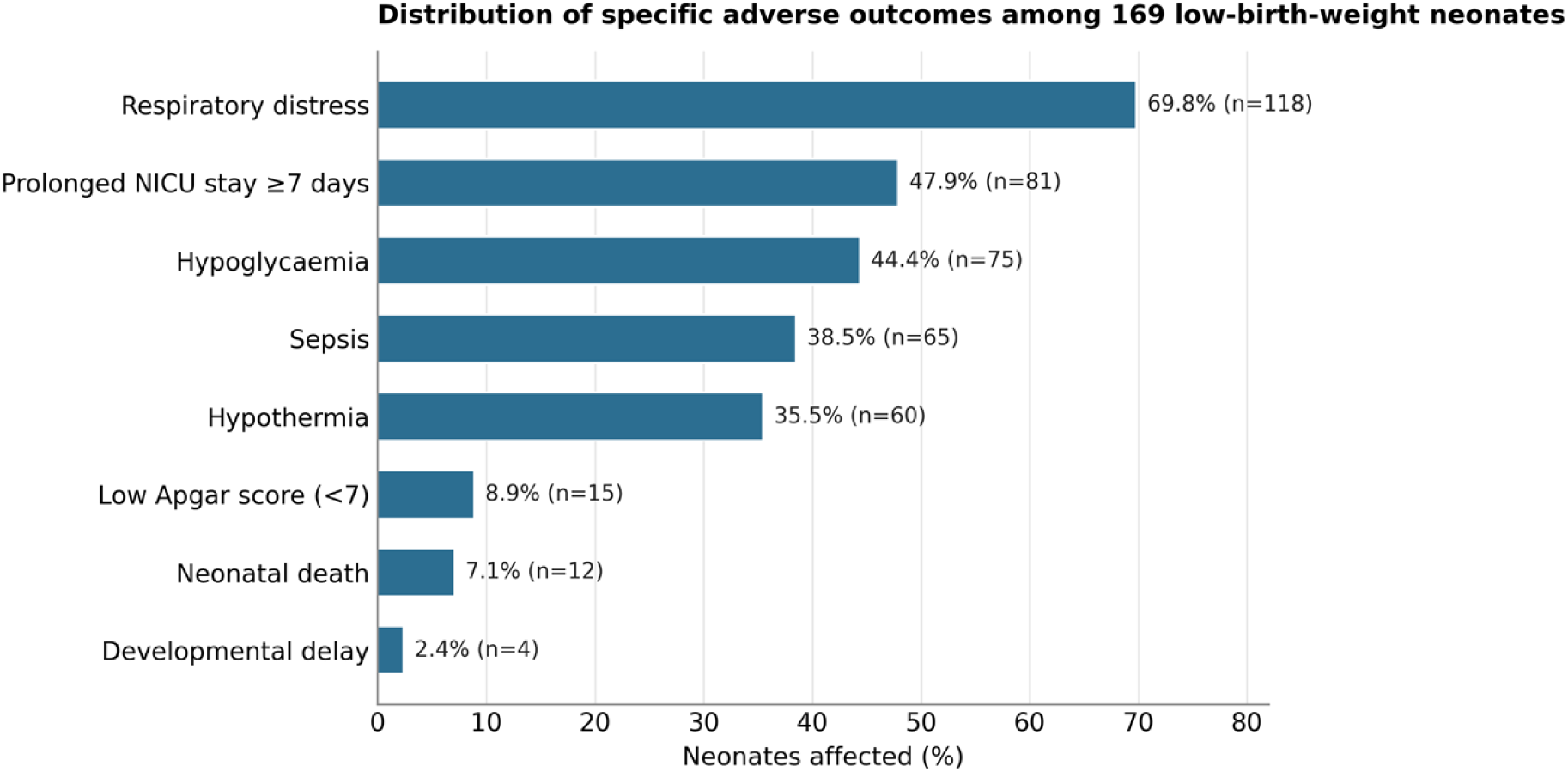
Distribution of specific adverse outcomes among 169 low-birth-weight neonates. Bars show the percentage (and number) of neonates affected by each complication; neonates could experience more than one outcome.

### Bivariate associations

At the bivariate level (Table 2), severe adverse outcome was significantly associated with maternal PIH (crude odds ratio [OR] 6.69, 95% CI 1.53–29.27, p = 0.004) and preterm birth (OR 3.79, 95% CI 1.50–9.56, p = 0.003). Neonates with a severe outcome had a substantially lower mean birth weight (1,704.8 vs 2,113.0 g, p < 0.001) and lower mean gestational age (31.9 vs 34.7 weeks, p < 0.001), and all 31 very-low-birth-weight neonates experienced a severe outcome (p = 0.002; crude OR not estimable owing to a zero cell). Antenatal-care attendance, congenital anomalies, sex, maternal age, parity and Apgar score were not significantly associated with severe adverse outcome.

**Table 2.** Bivariate associations between maternal and neonatal factors and severe adverse outcome.

| Variable | Severe (n = 136) | Not severe (n = 33) | $\chi^2 / t$ | Crude OR [95% CI] | p |
| --- | --- | --- | --- | --- | --- |
| Pregnancy-induced hypertension (PIH) | 41 (30.1%) | 2 (6.1%) | 8.12 | 6.69 [1.53, 29.27] | 0.004 |
| Attended ANC during pregnancy | 121 (89.0%) | 30 (90.9%) | 0.11 | 0.81 [0.22, 2.97] | 0.746 |
| Preterm birth | 122 (89.7%) | 23 (69.7%) | 8.73 | 3.79 [1.50, 9.56] | 0.003 |
| Congenital anomalies | 25 (18.4%) | 3 (9.1%) | 1.66 | 2.25 [0.64, 7.97] | 0.198 |
| Male sex | 64 (47.1%) | 20 (60.6%) | 1.95 | 1.73 [0.80, 3.76] | 0.163 |
| Very low birth weight (<1500 g) | 31 (22.8%) | 0 (0.0%) | 9.21 | — | 0.002 |
| Birth weight (g), M (SD) | 1704.8 (337.4) | 2113.0 (422.8) | 5.92 | — | <0.001 |
| Gestational age (weeks), M (SD) | 31.9 (4.0) | 34.7 (3.0) | 3.83 | — | <0.001 |
| Maternal age (years), M (SD) | 27.1 (6.7) | 28.3 (5.9) | 0.91 | — | 0.365 |
| Parity, M (SD) | 2.4 (3.2) | 2.2 (1.3) | -0.29 | — | 0.773 |
| Apgar score, M (SD) | 8.5 (1.5) | 8.5 (1.2) | -0.22 | — | 0.825 |

Note. Categorical variables are n (%); continuous variables are M (SD). $\chi^2$ = Pearson chi-square; t = independent-samples t test (Mann–Whitney U tests gave concordant results); OR = odds ratio; CI = confidence interval; ANC = antenatal care. The crude OR for very-low-birth-weight status was not estimable (zero cell).

### Multicollinearity diagnostics

Collinearity diagnostics (Table 3) showed acceptable values for all candidate predictors except the neonatal risk-factor count (VIF = 5.16), a composite of preterm birth, low birth weight, low Apgar score and congenital anomalies. Its adjusted estimate is therefore interpreted alongside its component variables rather than in isolation.

**Table 3.**
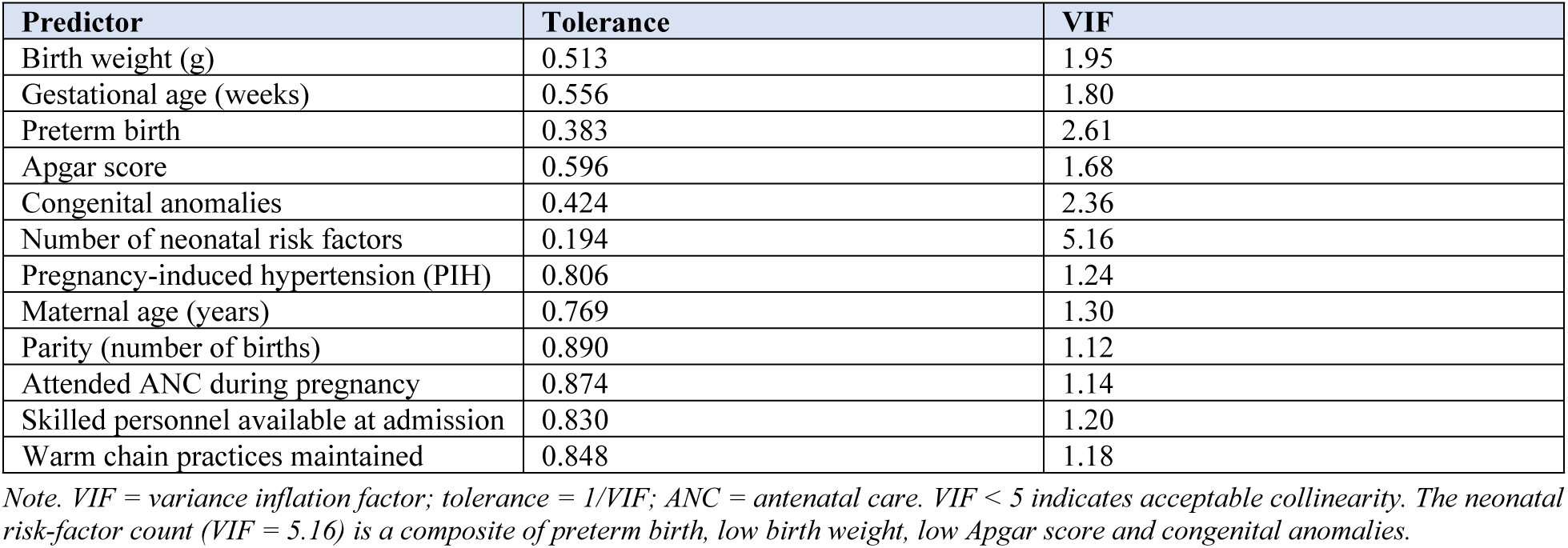
Multicollinearity diagnostics for candidate predictors.

| Predictor | Tolerance | VIF |
| --- | --- | --- |
| Birth weight (g) | 0.513 | 1.95 |
| Gestational age (weeks) | 0.556 | 1.80 |
| Preterm birth | 0.383 | 2.61 |
| Apgar score | 0.596 | 1.68 |
| Congenital anomalies | 0.424 | 2.36 |
| Number of neonatal risk factors | 0.194 | 5.16 |
| Pregnancy-induced hypertension (PIH) | 0.806 | 1.24 |
| Maternal age (years) | 0.769 | 1.30 |
| Parity (number of births) | 0.890 | 1.12 |
| Attended ANC during pregnancy | 0.874 | 1.14 |
| Skilled personnel available at admission | 0.830 | 1.20 |
| Warm chain practices maintained | 0.848 | 1.18 |

Note. VIF = variance inflation factor; tolerance = 1/VIF; ANC = antenatal care. VIF < 5 indicates acceptable collinearity. The neonatal risk-factor count (VIF = 5.16) is a composite of preterm birth, low birth weight, low Apgar score and congenital anomalies.

### Multivariable determinants of severe adverse outcome

The multivariable logistic regression model, entering all 13 candidate predictors simultaneously, was statistically significant (omnibus χ²(13) = 57.70, p < 0.001), well calibrated (Hosmer–Lemeshow χ²(8) = 4.45, p = 0.815), explained 46.1% of the variance in severe adverse outcome (Nagelkerke R² = 0.461), correctly classified 86.4% of cases and discriminated well (ROC area under the curve = 0.879, 95% CI 0.817–0.940). After simultaneous adjustment, three factors independently predicted severe adverse outcome (Table 4): lower birth weight (AOR 0.997 per gram, 95% CI 0.995–0.999, p = 0.012, indicating that the odds of a severe outcome fell as birth weight rose), maternal PIH (AOR 18.49, 95% CI 1.87–182.47, p = 0.013) and warm-chain care (AOR 10.94, 95% CI 1.40–85.41, p = 0.023).

**Table 4.**
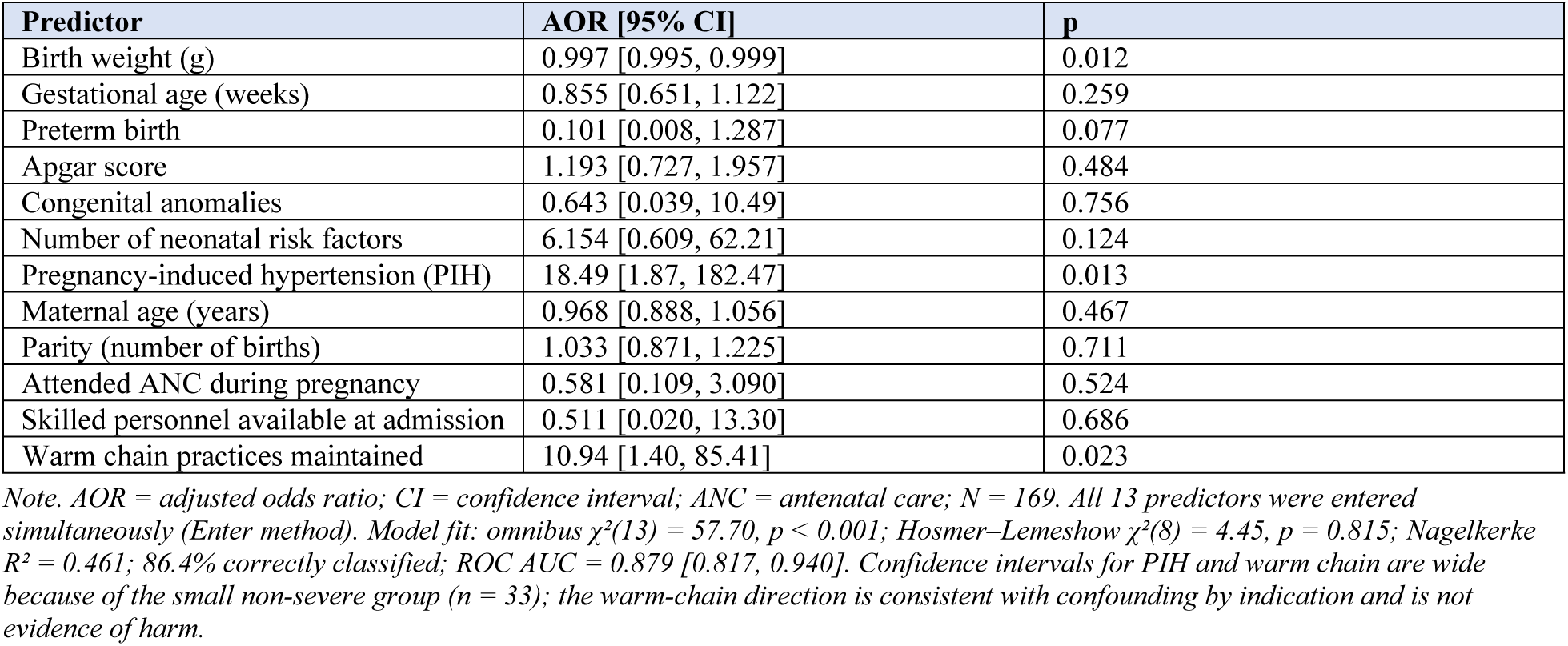
Multivariable logistic regression predicting severe adverse outcome (N = 169).

| Predictor | AOR [95% CI] | p |
| --- | --- | --- |
| Birth weight (g) | 0.997 [0.995, 0.999] | 0.012 |
| Gestational age (weeks) | 0.855 [0.651, 1.122] | 0.259 |
| Preterm birth | 0.101 [0.008, 1.287] | 0.077 |
| Apgar score | 1.193 [0.727, 1.957] | 0.484 |
| Congenital anomalies | 0.643 [0.039, 10.49] | 0.756 |
| Number of neonatal risk factors | 6.154 [0.609, 62.21] | 0.124 |
| Pregnancy-induced hypertension (PIH) | 18.49 [1.87, 182.47] | 0.013 |
| Maternal age (years) | 0.968 [0.888, 1.056] | 0.467 |
| Parity (number of births) | 1.033 [0.871, 1.225] | 0.711 |
| Attended ANC during pregnancy | 0.581 [0.109, 3.090] | 0.524 |
| Skilled personnel available at admission | 0.511 [0.020, 13.30] | 0.686 |
| Warm chain practices maintained | 10.94 [1.40, 85.41] | 0.023 |

*Note. AOR = adjusted odds ratio; CI = confidence interval; ANC = antenatal care; N = 169. All 13 predictors were entered simultaneously (Enter method). Model fit: omnibus $\chi^2(13) = 57.70$ , $p < 0.001$ ; Hosmer-Lemeshow $\chi^2(8) = 4.45$ , $p = 0.815$ ; Nagelkerke $R^2 = 0.461$ ; 86.4% correctly classified; ROC AUC = 0.879 [0.817, 0.940]. Confidence intervals for PIH and warm chain are wide because of the small non-severe group ( $n = 33$ ); the warm-chain direction is consistent with confounding by indication and is not evidence of harm.*

The wide confidence intervals for PIH and warm chain, and the counter-intuitive direction of the warm-chain association, reflect the small non-severe group (n = 33) and probable confounding by indication, whereby the smallest and most clinically unstable neonates were preferentially selected for active warm-chain care. These estimates are therefore interpreted as associations rather than causal effects, and the warm-chain finding should not be read as evidence that thermal care is harmful.

### Qualitative findings

Nine key informant interviews were conducted with one pediatrician, five nurses, two clinical officers and one newborn-unit manager, with 2–10 years of neonatal-care experience (Table 5). Thematic analysis generated eight interrelated care-related themes—timeliness of admission and triage, staffing and workload, warm-chain maintenance, infection-prevention practices, drug and equipment availability, referral coordination, monitoring and documentation, and caregiver delays and transport barriers (Table 6)—together with cross-cutting subthemes around the fragile “first hour” of care, pre-referral stabilisation and the role of mothers as co-providers of care through Kangaroo Mother Care and feeding support.

**Table 5.**
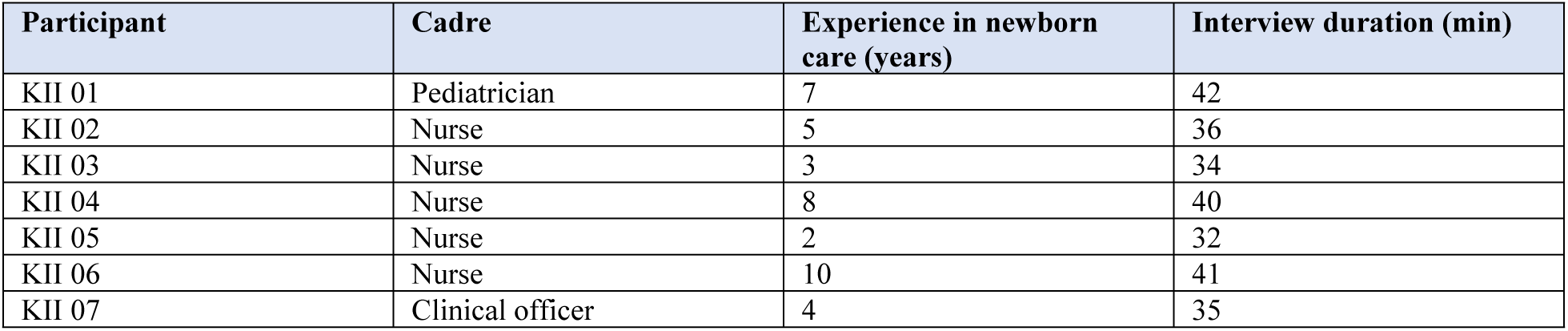
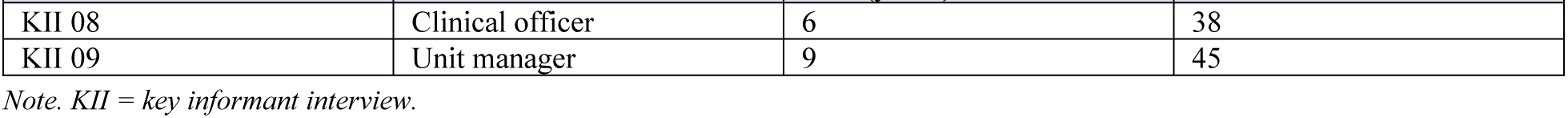
Characteristics of key informant interview participants.

| Participant | Cadre | Experience in newborn care (years) | Interview duration (min) |
| --- | --- | --- | --- |
| KII 01 | Pediatrician | 7 | 42 |
| KII 02 | Nurse | 5 | 36 |
| KII 03 | Nurse | 3 | 34 |
| KII 04 | Nurse | 8 | 40 |
| KII 05 | Nurse | 2 | 32 |
| KII 06 | Nurse | 10 | 41 |
| KII 07 | Clinical officer | 4 | 35 |
| KII 08 | Clinical officer | 6 | 38 |
| KII 09 | Unit manager | 9 | 45 |

Note. KII = key informant interview.

**Table 6.** Summary of qualitative themes on care-related determinants of adverse outcomes.

| Theme | Emerging subthemes | Illustrative meaning |
| --- | --- | --- |
| Timeliness of admission and triage | First hour of care; emergency stabilisation; congestion-related delays | Early assessment and intervention were viewed as critical for survival. |
| Staffing and workload | High nurse-to-baby ratio; night and weekend workload; burnout | Providers felt committed but overstretched. |
| Warm-chain maintenance | Kangaroo Mother Care dependence; insufficient incubators; cold referrals; unreliable power | Hypothermia was repeatedly linked to delayed or interrupted thermal care. |
| Infection-prevention practices | Congestion; caregiver movement; hand-hygiene and decontamination challenges; shared incubators | Infection prevention was difficult in a crowded unit. |
| Drug and equipment availability | Incubators, CPAP, oxygen, feeds, syringes, antibiotics | Supplies existed but were sometimes insufficient or inconsistent. |
| Referral coordination | Incomplete notes; lack of pre-arrival calls; poor pre-referral stabilisation; few ambulances | Referral gaps led to cold, unstable or poorly documented admissions. |
| Monitoring and documentation | Frequent observation needs; documentation sacrificed during emergencies | Inadequate monitoring increased the risk of missed deterioration. |
| Caregiver delays and transport barriers | Poor danger-sign recognition; family decision-making; cultural beliefs; cost and night transport | Delay often began before hospital arrival for referred neonates. |

Note. CPAP = continuous positive airway pressure. Themes were derived from nine key informant interviews with newborn-unit healthcare workers.

Providers consistently described the first hour after birth or arrival as decisive for LBW neonates, when delayed warming, oxygen support, glucose assessment or antibiotics could worsen prognosis. Staffing and workload were among the most prominent concerns; as one nurse explained, care could be slow because “there is low staffing which makes the care of the babies difficult in the facility from time of triage to admission of the babies is slow.” Warm-chain maintenance was repeatedly linked to hypothermia and to system constraints, including unreliable power: “Due to unreliable power supply within the facilities it makes the care for the babies difficult as incubators stop working hence babies start to lose heat.” Infection prevention was described as difficult in a crowded unit with intermittent supplies, with sepsis attributed partly to shared incubators and inconsistent decontamination. Commodity and equipment gaps—particularly shortages of continuous positive airway pressure (CPAP) machines and essential drugs—were said to delay management, and referral was viewed as poorly coordinated: “There are always delays for admission of babies… from other lower facilities since the referral system is not clear, [and the] mode of transport to the hospital also is not efficient as there are fewer ambulances in the county.” Caregiver-level delays were linked to limited danger-sign recognition, family decision-making and cultural beliefs, with some families interpreting LBW as “a curse or bad omen.” An overarching theme was that adverse outcomes reflected the interaction between clinical skill and system readiness.

Providers framed the earliest period of care as decisive. A pediatrician argued that newborn care should be treated as an emergency service, “because for these small babies, even one hour of delay can change the outcome” (KII 01), a view a clinical officer echoed in insisting that “the first hour after admission should be treated as critical” (KII 07). Competing admissions repeatedly eroded this window; as one nurse put it, “you may be stabilizing one baby on oxygen, then another baby arrives from theatre or maternity” (KII 05).

Staffing pressure was tied directly to monitoring and documentation. A nurse observed that during emergencies “documentation suffers because the priority becomes keeping the baby alive first” (KII 02), while a more experienced colleague cautioned that “you cannot manage these babies like ordinary patients because small changes matter” (KII 04). The unit manager conceded that although staff were skilled, “burnout is real” (KII 09).

Thermal care, equipment and crowding were closely intertwined. When incubators were full, staff improvised—“a stable baby may have to be moved to KMC earlier so that a more critical baby can use an incubator” (KII 03)—and congestion undermined infection control, because “when the unit is crowded, it becomes difficult to separate infected and non-infected babies properly” (KII 04).

Referral gaps produced cold, undocumented arrivals. A pediatrician described babies “referred on motorbikes or public vehicles, wrapped in ordinary clothes, with no oxygen or warmth,” so that “by the time they arrive, the situation is already worse” (KII 01), and a clinical officer noted that such neonates often “arrive with several problems at once: cold, hypoglycemic, and breathing poorly” (KII 08). Referral notes were frequently too sparse to guide management—one nurse received a note that “only [said] ‘premature baby’ without temperature, blood sugar, treatment given, or Apgar score,” which “makes us repeat assessment and lose time” (KII 02)—leading the unit manager to insist that “a small baby should not be transported cold” (KII 09).

Delays often began before admission. Providers attributed these to weak danger-sign recognition—“poor feeding, fast breathing, and cold skin may not be taken seriously early” (KII 08)—and to family decision-making and cost, with families delaying “because they do not know danger signs or they are looking for money” (KII 04). The mother’s own state then shaped care on the ward: warm-chain maintenance was easier “when the mother understands KMC,” but “if the mother is tired, post-operative, or afraid to hold a small baby, it becomes difficult” (KII 03).

### Integration of quantitative and qualitative findings

The joint display (Table 7) shows convergence between the two strands. The near-universal burden of respiratory and thermal-metabolic complications aligned with provider accounts of a fragile first-hour stabilisation window; the independent effect of birth weight matched providers’ emphasis on the intrinsic vulnerability of the smallest neonates; the independent maternal-PIH effect was consistent with provider observations linking maternal antenatal conditions to fragile newborns; and the warm-chain and system themes explained why thermal and care-readiness variables were statistically entangled with illness severity. Together, the strands indicate that neonatal vulnerability and maternal hypertensive disease set the level of risk, while the readiness and timeliness of newborn-unit care shaped whether that risk translated into a severe outcome. This reading was crystallised by the unit manager, who observed that “staff can do their best, but without enough personnel, equipment, and referral coordination, some preventable complications will continue” (KII 09).

**Table 7.** Joint display of quantitative and qualitative findings by objective.

| Objective | Key quantitative finding | Supporting qualitative theme | Integrated interpretation |
| --- | --- | --- | --- |
| Types of adverse outcomes | Adverse outcomes were near-universal: 160 (94.7%) had $\geq 1$ and 136 (80.5%) a severe outcome; respiratory distress most common (69.8%); mean 2.46 (SD 1.31) per neonate. | Providers described the fragile “first hour” of warming, oxygen, glucose monitoring and early feeding as decisive. | Most LBW neonates experienced multiple, co-occurring complications dominated by respiratory and thermal instability, consistent with a fragile early-stabilisation window. |
| Maternal determinants | PIH was associated with severe outcome at bivariate level (OR 6.69, $p = 0.004$) and remained an independent predictor after adjustment (AOR 18.49, $p = 0.013$). | Providers linked poor antenatal preparation and maternal illness to fragile newborns and to reduced capacity to sustain Kangaroo Mother Care and feeding. | Maternal hypertensive disease independently raised the odds of a severe outcome, acting both through and beyond neonatal size. |
| Neonatal determinants | Lower birth weight, lower gestational age and preterm birth were significant at bivariate level; birth weight was an independent predictor after adjustment (AOR 0.997/g, $p = 0.012$). | Providers emphasised the intrinsic vulnerability of very small and preterm babies requiring frequent monitoring and rapid escalation. | Neonatal immaturity, best summarised by birth weight, was the most consistent determinant of severe adverse outcome. |
| Care-related determinants | Warm-chain care was an independent predictor in a direction consistent with confounding by indication (AOR 10.94, $p = 0.023$); other care variables were not independent predictors. | Providers described staffing shortages, intermittent equipment and supplies, unreliable power, weak referral coordination and caregiver or transport delays. | Care-related factors were entangled with illness severity but remained actionable, system-level levers for improving outcomes. |

*Note. AOR = adjusted odds ratio; OR = odds ratio; PIH = pregnancy-induced hypertension; LBW = low birth weight.*

## Discussion

### Principal findings

In this mixed-methods study of 169 LBW neonates admitted to a Kenyan county referral hospital, severe adverse outcomes were near-universal (80.5%), and most neonates experienced multiple, co-occurring complications dominated by respiratory distress and thermal-metabolic instability. After simultaneous adjustment, three factors independently predicted a severe adverse outcome—lower birth weight, maternal pregnancy-induced hypertension and warm-chain care—in a model with strong discrimination (AUC 0.879). Provider narratives explained these associations by locating neonatal vulnerability within a care system whose timeliness, staffing, thermal care, supplies and referral coordination determined whether biological risk progressed to a severe outcome.

### Burden and pattern of adverse outcomes

The high burden of respiratory distress, prolonged admission, hypoglycaemia, sepsis and hypothermia is consistent with the known physiological vulnerability of LBW and preterm neonates and with evidence from comparable settings [6, 12, 26]. Respiratory distress predominated, reflecting pulmonary immaturity and the referral role of KCRH, which concentrates the most severe presentations. The frequency of hypoglycaemia and sepsis underscores the importance of early feeding and glucose surveillance and of timely infection prevention and antibiotic administration, since the newborn brain is highly vulnerable to glucose deprivation and neonatal sepsis remains a leading cause of morbidity and mortality [8, 9, 11]. Hypothermia, present in a third of neonates, is clinically important because it can worsen respiratory distress, hypoglycaemia and sepsis, and Kenyan evidence links it to LBW and to increased mortality [10, 27]. The observed clustering of complications supports treating admitted LBW neonates as a high-risk group requiring structured assessment, monitoring and early stabilisation [28, 29].

### Neonatal vulnerability and birth weight

Birth weight was an independent predictor of severe adverse outcome, with each additional gram associated with lower odds—equivalent to a meaningful reduction per 100 g. Lower birth weight captures reduced physiological reserve, immature organ function and limited metabolic stores, which help explain why it retained significance alongside maternal and care-related factors. This finding is consistent with African and global evidence that lower birth weight and prematurity are strongly associated with neonatal morbidity and mortality [30–32]. The near-perfect separation observed for very-low-birth-weight status—every such neonate had a severe outcome—reinforces birth weight as both a clinical and a programmatic risk marker that can be used at admission to identify neonates requiring intensified monitoring and early intervention [5].

### Maternal pregnancy-induced hypertension

Maternal PIH was independently associated with severe adverse outcome, a finding that is biologically plausible: hypertensive disorders impair uteroplacental perfusion and are linked to placental insufficiency, fetal growth restriction and preterm birth, producing neonates who are smaller and less physiologically prepared for extrauterine life [33, 34]. That PIH persisted as an independent predictor—rather than being fully mediated by birth weight—suggests that hypertensive disease may exert effects on neonatal stability beyond size alone, for example through the circumstances of delivery and the condition of the infant at presentation. The wide confidence interval reflects the small number of non-severe cases and warrants cautious interpretation, but the direction is consistent with the wider literature on maternal hypertensive disease and adverse neonatal outcomes [16, 34]. The finding strengthens the case for antenatal blood-pressure screening, early detection and management of hypertensive disorders, and timely referral of affected pregnancies [35, 36].

### Warm-chain care, care-related factors and confounding by indication

Warm-chain care emerged as an independent predictor, but in a counter-intuitive direction that almost certainly reflects confounding by indication: the smallest, most unstable neonates were preferentially placed on active thermal support, so the marker of receiving warm-chain care became entangled with illness severity. This interpretation is reinforced by the qualitative data, in which providers uniformly regarded warm-chain maintenance as essential and described hypothermia as a consequence of cold referrals, delayed skin-to-skin contact and unreliable power rather than of thermal care itself. Established evidence that neonatal hypothermia is common, is associated with LBW and increases mortality, and that adherence to thermal-care guidelines is often incomplete in Kenyan units, argues strongly against reading this association as evidence of harm [10, 37, 38]. More broadly, care-related factors—staffing and workload, commodity and equipment availability, infection prevention and referral coordination—were emphasised by providers as modifiable determinants even where they were not independent statistical predictors, echoing evidence that NICU staffing and work environment influence infection rates and quality of care and that reliable implementation is essential for evidence-based neonatal interventions [39–43].

### Referral, delays and interpretation within the theoretical frameworks

Provider accounts of cold, undocumented and delayed referrals, limited ambulance availability and caregiver delays map directly onto the Three Delays Model. Delay 1 was reflected in limited caregiver recognition of newborn danger signs and family decision-making, including cultural beliefs about small babies [44, 45]; Delay 2 in transport barriers, distance and weak referral coordination [46–48]; and Delay 3 in triage, staffing, supplies and the fragile first hour after arrival [49, 50]. The Social Ecological Model situates these findings across levels: neonatal biological vulnerability and maternal hypertensive disease at the individual level; family decision-making and Kangaroo Mother Care at the interpersonal level; residence, distance and transport at the community level; and staffing, commodities, referral systems and documentation at the institutional and policy levels [18, 51–54]. Together the frameworks support the central message that severe adverse outcomes were driven by neonatal and maternal risk but shaped by the readiness of the care pathway.

### Implications for practice and policy

The findings support birth-weight-based risk stratification at admission, with the smallest neonates prioritised for intensified monitoring, thermal protection, respiratory assessment, glucose surveillance, feeding support, infection prevention and early escalation [29, 55]. The independent maternal-PIH effect reinforces the value of antenatal detection and management of hypertensive disease and of anticipatory newborn-unit preparation for affected pregnancies. At facility and county level, the provider-identified constraints point to actionable levers: adequate newborn-unit staffing, reliable commodities and functional equipment (including CPAP and a dependable power supply), strengthened warm-chain and infection-prevention practices, structured monitoring and documentation, and improved pre-referral stabilisation and referral communication, consistent with national newborn-care protocols [56, 57].

### Strengths and limitations

Strengths include the locally relevant, facility-specific focus; the mixed-methods design, which paired measured associations with provider explanations; the use of the full cohort in the adjusted model; and the reporting of calibration, classification and discrimination diagnostics. The study also used a clinically meaningful severe-outcome variable in place of a near-universal composite, improving analytic interpretability.

Several limitations should temper interpretation. The cross-sectional, single-site design precludes causal inference and limits generalisability, and a referral hospital concentrates severe presentations. The outcome distribution was imbalanced, with only 33 non-severe cases; entering 13 predictors in this setting risks overfitting, and the wide confidence intervals for PIH and warm chain reflect this, so these estimates should be regarded as hypothesis-generating. Warm-chain care and neonatal-unit admission are susceptible to confounding by indication and should not be interpreted causally. Some exposures relied on maternal report and were subject to recall and classification error, and several conceptually relevant variables were incompletely captured, raising the possibility of residual confounding. Larger, multi-site, prospective studies with stronger measurement of care processes are needed to confirm these determinants.

**Fig 2.**
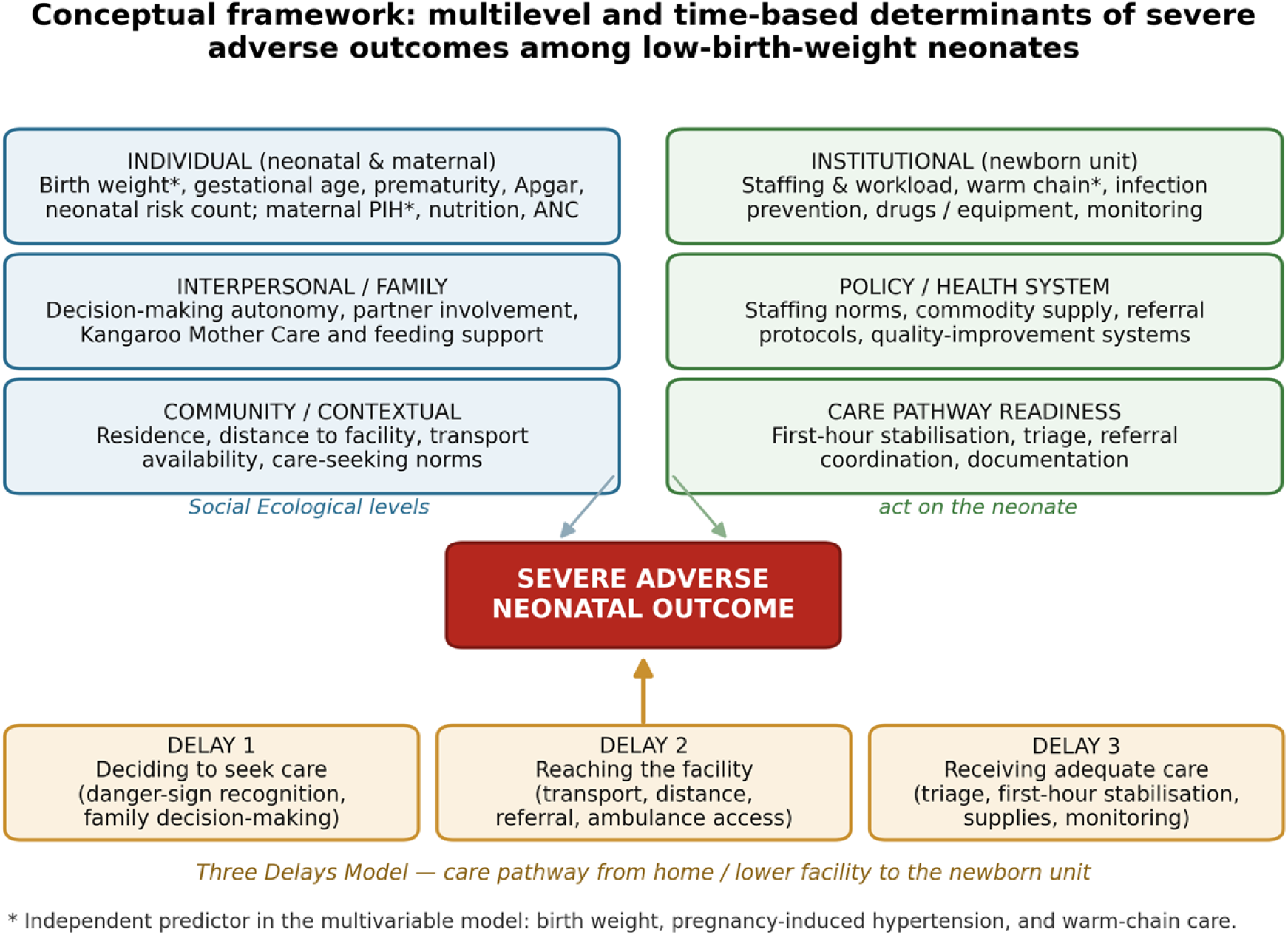
Conceptual framework integrating the Social Ecological Model and the Three Delays Model. Individual, interpersonal, community, institutional and policy-level determinants interact with delays in deciding to seek care, reaching the facility and receiving adequate care to shape severe adverse neonatal outcomes. Asterisks denote factors that were independent predictors in the multivariable model (birth weight, pregnancy-induced hypertension and warm-chain care).

## Conclusions

Severe adverse outcomes were near-universal among LBW neonates admitted to Kericho County Referral Hospital and were driven by neonatal biological vulnerability, maternal pregnancy-induced hypertension and the readiness of newborn-unit care. Birth weight was a robust, independent marker of risk, maternal hypertensive disease independently predicted severe outcome, and the warm-chain association is best understood as confounding by indication within a strained care system. Improving outcomes for this population requires early, birth-weight-based identification of the most vulnerable neonates, strengthened antenatal detection and management of hypertensive disease, and reliable, timely newborn-unit care processes—adequate staffing, dependable commodities and equipment, consistent warm-chain and infection-prevention practice, and better pre-referral stabilisation and referral coordination.

## Acknowledgements

The authors thank the mothers and healthcare workers who participated in the study and the management and newborn-unit staff of Kericho County Referral Hospital for their support during data collection.

## Author contributions

JC conceived and designed the study, collected the data and drafted the manuscript. MSS contributed to study design, supervised the analysis and critically revised the manuscript. MK contributed to study design and supervision. EN contributed to the analysis and interpretation of the data. All authors read and approved the final manuscript.

## Data availability

The de-identified data supporting the findings of this study are available from the corresponding author on reasonable request, subject to the ethical-approval conditions governing the study.

## Ethics statement

Ethical approval was obtained from the Kabarak University Institutional Scientific and Ethics Review Committee and the National Commission for Science, Technology and Innovation (NACOSTI), with administrative authorisation from Kericho County and Kericho County Referral Hospital. Written informed consent was obtained from all participants.

